# *STARD9* and *CDK5RAP2* – novel candidate genes for oligogenic 46,XY complete gonadal dysgenesis

**DOI:** 10.1101/2025.05.10.25326049

**Authors:** Dmytro Sirokha, Alexey Rayevsky, Vitalii Kalynovskyi, Mykola Khalangot, Oksana Samson, Olexandra Gorodna, Krystyna Kwiatkowska, Zaneta Lemanska, Amanda Kunik, Chloe Mayere, Serge Nef, Kamila Kusz-Zamelczyk, Ludmila Livshits

## Abstract

46,XY gonadal dysgenesis (46,XY GD) results from disruptions in the genetic program that governs testicular differentiation during gonadal sex determination, presenting as either complete (46,XY CGD) or partial (46,XY PGD) forms. While monogenic defects account for approximately 50% of cases, recent evidence suggests an oligogenic basis for some 46,XY GD cases. In this study, we investigated a case of 46,XY CGD and performed whole-exome sequencing (WES) on the patient and her parents to explore the genetic basis of the patient’s condition. Although no pathogenic variants were identified in known 46,XY GD-associated genes, we detected rare variants in the *STARD9* and *CDK5RAP2* genes. Previous study in mice indicate that the orthologues of these genes are highly expressed in Sertoli cells during gonadal sex determination, with *Cdk5rap2* playing a critical role in Sertoli cell polarization. Notably, the human STARD9 and CDK5RAP2 proteins interact with each other. Structural analysis suggests that the variants in STARD9 and CDK5RAP2 may alter their protein-protein interactions. Based on these findings, we propose that *STARD9* and *CDK5RAP2* variants may act together to impair Sertoli cell function, leading to 46,XY CGD, consistent with an oligogenic mode of inheritance. These results suggest that *STARD9* and *CDK5RAP2* should be considered as candidate genes for 46,XY GD and included in genetic panels for this condition.

## INTRODUCTION

Sexual development in humans depends on the proper determination, differentiation and functioning of the gonads. Genetic mutations and genomic rearrangements affecting these processes can lead to Disorders/Differences of Sex Development (DSD) where an individual’s chromosomal sex (XX or XY) does not align with their gonadal (ovary or testis) or phenotypical sex (1). In the case of male chromosomal sex, disruption of genetic program that controls testicular development leads to 46,XY gonadal dysgenesis (46,XY GD). This condition encompasses 46,XY complete GD (46,XY CGD) characterized by complete absence of testicular tissue resulting in female genitalia and 46,XY partial GD (46,XY PGD) characterised by impaired development of testicular tissue resulting in genital ambiguity (2).

Approximately 50% of 46,XY GD cases are attributed to mutations in genes such as *SRY, NR5A1, MAP3K1*, and *DHX37*, while mutations in other 46,XY GD genes are very rare. Recently, WES allowed the identification of several new candidate genes, yet the genetic basis of nearly 50% of 46,XY GD cases remains unresolved (for a comprehensive review, refer to (2)). Increasing evidence suggests that some cases follow an oligogenic inheritance pattern, where multiple genetic variants collectively contribute to the phenotype (3-5).

In this study, we describe a 46,XY CGD patient in whom WES did not reveal pathogenic variants in known 46,XY GD-associated genes. Instead, we detected variants in *Steroidogenic Acute Regulatory-Related Lipid Transfer Domain 9* (*STARD9*) and *CDK5 Regulatory Subunit-Associated Protein 2* (*CDK5RAP2*), two genes encoding proteins that interact with each other (6). Given that previous studies in mice indicate that orthologues of these genes are highly expressed in Sertoli cells during gonadal sex determination (7), with *Cdk5rap2* playing a critical role in Sertoli cell development (8), we propose that the combined effect of *STARD9* and *CDK5RAP2* variants may contribute to 46,XY CGD, supporting an oligogenic mode of inheritance.

## MATERIALS AND METHODS

### The patient

The patient – UKR21 was born after the first normal pregnancy to healthy non-consanguineous parents, both between the ages of 24 to 28. The birth weight was 3500 g and the length was 55 cm. At birth, the child was registered as a female. At adolescence age, the patient visited a gynaecologist due to an absence of puberty signs and lack of breast development.

### Hormonal analysis

Levels of free testosterone (fT), luteinizing hormone (LH), follicle-stimulating hormone (FSH), oestradiol (E2) and progesterone (P4) in serum were quantified using electrochemiluminescence immunoassay technology on the Cobas E411 analyser (Roche Diagnostics, Risch-Rotkreuz, Switzerland). The assays were performed using the Elecsys Testosterone II, LH, FSH, E2 III, and Progesterone III kits, following the manufacturer’s instructions (Roche Diagnostics, Mannheim, Germany).

### Cytogenetic studies

Cytogenetic studies were performed on peripheral blood lymphocytes (30 metaphase plates) using Nikon Eclipse Ci microscope (Nikon, Minato, Japan). FISH analysis was performed at 200 interphase nuclei using LUCIA Cytogenetics Software (Praha, Czech Republic) according to Cytogenetic Guidelines and Quality Assurance by European Cytogenetics Association (GTG-banding, FISH-probes CEP, LSI (probes: Yp11.3 - *SRY*; Yp11.1-q11.1 - *DYZ3*; Yq12 - *DYZ1*; CEP - *DXZ1*)) (Abbott Molecular, Libertyville, IL, USA).

### Histologic studies

Gonad sections were taken following gonadectomy. The sections were fixed in neutral buffered formalin for 72 hours, then dehydrated by three changes of 99% isopropanol, cleared in xylene, and embedded in paraffin. Morphological analysis was performed on 5-μm sections that were routinely stained with Ehrlich haematoxylin and eosin. Samples were analysed using light microscopy with ×40–×100 magnification.

### Bone tissue and brain studies

Bone tissue studies included densitometry, which was performed using a Discovery Wi bone densitometer (Hologic, Marlborough, MA, USA) and TBS iNsight™ software (Medimaps Group, Plan-les-Ouates, Switzerland). Brain studies included magnetic resonance imaging, which was performed using the Symphony 1.5 Tesla MRI system (Siemens, Munich, Germany).

### Whole-exome sequencing (WES)

Genomic DNA (gDNA) from the blood samples of the patient and her parents was isolated by using the QIAmp DNA Kit (Qiagen, Hilden, Germany). Exome capture was performed on the DNA samples using the SureSelectXT Target Enrichment system for Illumina version B.2 (Agilent Inc®, Santa Clara, CA, USA). Paired-end libraries were prepared using TruSeq SBS Kit v3 (Illumina, San Diego, CA, USA) and sequenced on an Illumina HiSeq 4000 system (Illumina, San Diego, CA, USA). Raw sequencing was transformed into .fastq files using the CASAVA v1.8.1 software (Illumina, San Diego, CA, USA) and processed with DRAGEN Germline Pipeline v 2.3 (Edico Genome), which leverages Genome Analysis Tool Kit (GATK; https://gatk.broadinstitute.org/hc/en-us/articles/360045944831) and is harboured at the Illumina’s cloud-based resource BaseSpace.

For the proband and her parents, 100 to 150 million reads were processed, adapter trimmed, duplicate marked, and aligned against the GRCh37/hg19 assembly of the human genome using Smith-Waterman scoring algorithm. At the variant calling stage, the following filters were applied: variant confidence/quality by depth > 2.0, mapping quality > 30.0, phred-scaled p-value for strand bias < 60.0, mapping quality RankSum > 12.5, ReadPos-RankSum > 8.0. Variants were annotated and analysed in Variant Interpreter (Illumina, San Diego, CA, USA) and VarSeq (Golden Helix, Boseman, MT, USA). Further filtering was performed by the variant quality > 500, genotyping quality > 80, read depth > 30, proportion of reads bearing the minor allele > 0.2, and population frequency < 0.01.

### Validation of the *STARD9* and *CDK5RAP2* variants by Sanger sequencing

The status of the *STARD9*(NM_020759.3):c.3514C>T (p.Arg1172Cys), the *STARD9*(NM_020759.3):c.5585_5590del (p.Ser1862_Thr1863del), and the *CDK5RAP2*(NM_018249.6):c.2003A>G (p.Tyr668Cys) variants in the patient and her parents was confirmed by Sanger sequencing. DNA fragments, each containing one of the above variants, were amplified using the appropriate gDNA templates and specific primers (forward primer TTTCCCAGAGCCAGAGAACT and reverse primer CCCAGTTCTTCCTCTGCAT for the *STARD9:*c.3514C>T variant; forward primer CCTCATCTCAGCAGGTCACA and reverse primer TCCTCTCGTGCCTCAGATTC for the *STARD9:*c.5585_5590del variant; forward primer CCTGGGAAGCTGAAGTCTCT and reverse primer CGCAAGTCTATCTGGAAACCC for the *CDK5RAP2:*c.2003A>G variant). Amplification was performed using the Advantage® 2 PCR Enzyme System with Advantage 2 PCR Buffer or Advantage 2 PCR SA Buffer (Takara Bio, San Jose, CA, USA) according to the manufacturer’s instructions. Each amplicon was sequenced bidirectionally using the respective primers in separate reactions. Additionally, the *STARD9* fragment containing the c.5585_5590del variant was cloned to the pGEM-T Easy vector and resequenced to obtain readable chromatograms. Sequencing was performed using BigDye™ Terminator v3.1 Cycle Sequencing Kit on a 3730xl DNA Analyzer (Applied Biosystems, Waltham, MA, USA). Chromatograms were analysed using SnapGene 4.3.11 software (GSL Biotech LLC, San Diego, CA, USA).

### Molecular modelling and molecular dynamics (MD) simulations

To determine the secondary structure of the wild-type STARD9 fragment containing residues Arg1172 and Ser1862_Thr1863 (which are substituted or deleted, respectively, in the patient) several disorder predictors were used: FoldIndex (9), NORSp (10), GlobPlot (11), and CH-Plot (12). These predictors rely on the physical and chemical characteristics of amino acids and data from known unstructured proteins. Additionally, IUPred (13), Ucon (14) and FoldUnfold (15) inter-residue contact-based predictors were applied. The three-dimensional structures were constructed using AlphaFold2 (16). Furthermore, the coarse-grained MD simulation of the wild-type STARD9’s large central segment (1136-1888 aa) containing the Arg1172 and Ser1862_Thr1863 residues flanked by 25-36 residues was analysed using the Gromacs 2018.1 software tool (17), along with the CHARMM36 (18) and SIRAH2.0 (19) force fields. The simulation was continued for 700 ns. A constant system temperature was maintained using a Nosé-Hoover thermostat, with simulations performed at 310K and 1 bar pressure, controlled by a Parrinello-Rahman barostat. A cut-off radius of 12 Å was applied for both Coulomb (electrostatic) and Lennard-Jones (van der Waals) interactions. The Particle-Mesh Ewald method for long-range interactions was used with periodic boundary conditions in the XYZ directions. Finally, the full-atom MD simulation of two mutated STARD9’s short fragments (1163-1179 aa and 1849-1872 aa), each containing either the p.Arg1172Cys or p.Ser1862_Thr1863del mutation flanked by 7-13 residues, was performed for 500 ns to induce secondary structure ordering. Define secondary structure of proteins analysis (20) was applied to assess the propensity of these structures to fold under near-natural conditions, and the results were compared with predicted secondary structure assignments.

The CDK5RAP2 protein was rebuilt using AlphaFold2 server (16). MD of a single helix (650-684 aa that contains Tyr668 residue mutated in the patient) was performed using the Gromacs 2018.1 software tool (17), along with the CHARMM36 (18). The simulation was continued for 200 ns, under the same conditions, that we used for the simulations of STARD9 protein fragments.

### Bioinformatic resources and tools

The Genome Aggregation Database v4.1.0 (https://gnomad.broadinstitute.org/) was used to check the allele frequency of the variants. The public medical genetics databases ClinVar (https://www.ncbi.nlm.nih.gov/clinvar/) and VarSome (https://varsome.com/) were used to check if the variants were registered in any disease. The BioGRID (https://thebiogrid.org/) and IntAct (https://www.ebi.ac.uk/intact/home) protein-protein interaction databases were used to find STARD9’s protein interactors.

### Accession numbers

The following accession numbers for transcripts were used: NM_020759.3 for *Homo sapiens STARD9*, NM_018249.6 for *Homo sapiens CDK5RAP2*. The following accession numbers for mammalian STARD9 proteins were used: Q9P2P6 for *Homo sapiens*, G3QVK0 for *Gorilla gorilla*, XP_020952823.1 for *Sus scrofa*, F6W6H9 for *Equus caballus*, and Q80TF6 for *Mus musculus*. The following accession numbers for mammalian CDK5RAP2 proteins were used: Q96SN8 for *Homo sapiens*, G3QD67 for *Gorilla gorilla*, A0A8W4FHP0 for *Sus scrofa*, F7DTT1 for *Equus caballus*, and Q8K389 for *Mus musculus*.

## RESULTS

### The 46,XY CGD patient

At adolescence age, the female patient was referred to a gynaecologist due to the absence of puberty signs. Clinical assessment showed Tanner stages 1-2 breast development and Tanner stage 4 pubic hair. Pelvic ultrasonography revealed a 22 × 13 × 25 mm uterus, a 17 × 12 mm right gonad, and an 18 × 13 mm left gonad. Hormonal investigations revealed low oestradiol (E2 31.94 pmol/L), low progesterone (P4 1.65 nmol/L), and low free testosterone (fT 0.7628 nmol/L), while gonadotropin levels were markedly elevated (FSH 153.2 IU/L, LH 41.8 IU/L), consistent with primary (hypergonadotropic) hypogonadism. Cytogenetic analysis revealed a 46,XY karyotype with the presence of a *SRY* gene and no mosaicism. Based on the presence of female external genitalia and uterus, small gonads, primary amenorrhea, elevated gonadotropins, and a 46,XY karyotype, the diagnosis of 46,XY CGD was made.

Shortly afterwards, the patient underwent laparoscopic gonadectomy, which revealed whitish gonads at the ends of the rudimentary fallopian tubes. Both the gonads and fallopian tubes were excised. Histological analysis confirmed complete gonadal dysgenesis, with the gonads composed of fibrous connective tissue and no differentiated structures observed (**Fig. 1**). The cortical and medullary layers were distinguishable, with the cortical layer containing single follicular structures and detached epithelial tissue (**Fig. 1a** and **b**), while the medullary layer was characterized by numerous blood vessels and lacunae (**Fig. 1c**). Additionally, a thickened fallopian tube with a highly branched lumen was observed (**Fig. 1d**). Following sex hormone replacement therapy, the patient experienced regular menstrual cycles.

**Fig. 1.**
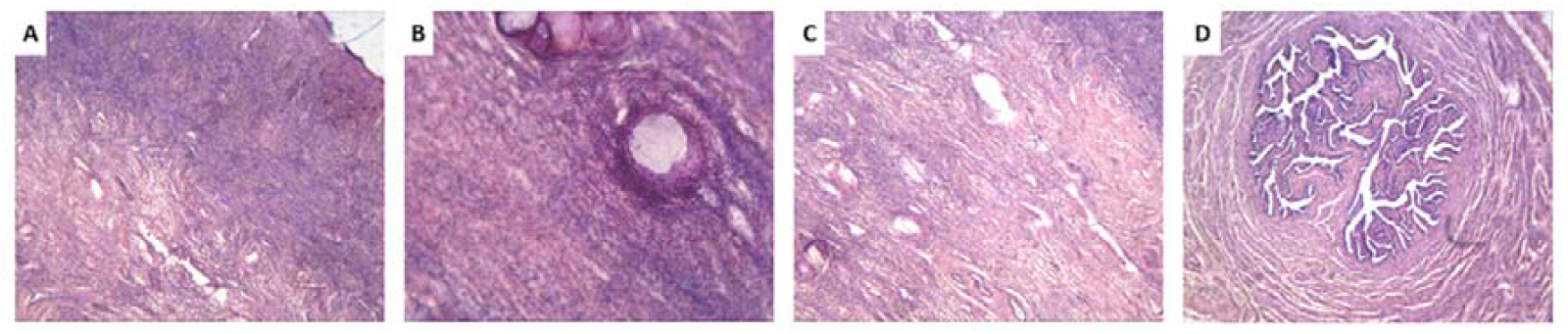
The histology of the patient’s gonad, stained with haematoxylin and eosin. **A.** The gonad (only one gonad was documented) formed by fibrous connective tissue. The cortical layer of the gonad is more dense then the medullar layer. Magnification x40. **B**. The cortical layer with single follicular-like structures with detached epithelial tissue and homogeneously faintly stained basophilic substance. Magnification x100. **C**. The medullar layer with numerous blood vessels and lacunae. Magnification x40. **D**. Oviduct. Magnification x40.

In addition to 46,XY CGD, the patient exhibited delayed bone age; at 15-19 years old, her bone age corresponded to 12-13 years. At that time, her height was 167 cm, and weight was 48 kg. At the age of 26-30, she had grown to 172 cm, and weighted 53 kg. At the age of 15-19, she was diagnosed with osteoporosis (z-score: -2.4 at the femoral neck, -2.5 at L1–L4, and -0.9 at the radius), which progressed to systemic osteoporosis by age 28 (t-score -2.5, z-score -2.5). The early onset of osteoporosis was likely due to lack of gonadal hormone secretion and delayed initiation of oestrogen replacement therapy. Brain magnetic resonance imaging revealed no abnormalities. The final diagnosis for the patient was 46,XY CGD and osteoporosis.

### WES outcome

To identify the genetic cause of 46,XY CGD in the patient, WES was performed on both the patient and her parents. The sequencing achieved a mean coverage of > 145, with 95.3% of the targets covered at depth of > 30. Initial analysis revealed 1’360’189 single nucleotide variants (SNV) or small insertion-deletion (indel) variants in the patient. After filtering based on quality and population frequency, a subset of 509 coding sequence variants was retained for further analysis.

### Variants in known DSD genes

We first screened these 509 variants against a curated list of 194 genes associated with DSD including known and candidate genes (21-30).

We identified three heterozygous variants in autosomal DSD candidate genes: *Chromodomain Helicase DNA Binding Protein 7* (*CHD7*) NM_017780.4:c.2273G>A (p.Arg758His) (rs202208393), *KISS1 Receptor* (*KISS1R*) NM_032551.5:c.1167C>A (p.Cys389Ter) (rs371771794), and *Leucine Rich Repeat Containing G Protein-Coupled Receptor 5* (*LGR5*) NM_003667.4:c.2341C>G (p.Pro781Ala) (rs113809442). However, segregation analysis revealed that all three variants were inherited from the father, who did not exhibit any DSD features, suggesting that these variants alone are not sufficient to cause 46,XY CGD. Nevertheless, we cannot exclude their potential modifying effect on our patient’s phenotype, particularly since CHD7 has been shown to directly regulate *Sry* expression in mice (31).

### *STARD9* variants in a compound heterozygous state

Since no convincing pathogenic variants were identified in known DSD-related genes, we extended our analysis to other genes. Among the genes with rare variants detected in the patient, *STARD9* gene, which encodes a protein belonging to the STAR family, emerged as a strong candidate, as mutations in other STAR family genes have previously been implicated in 46,XY DSD (26, 29, 32). Furthermore, in mice, *Stard9* is expressed in a sex-specific manner during gonadal differentiation, with significantly higher expression in Sertoli cells compared to granulosa cells, supporting its potential role in testicular development (**Fig. 2**) (7).

**Fig. 2.**
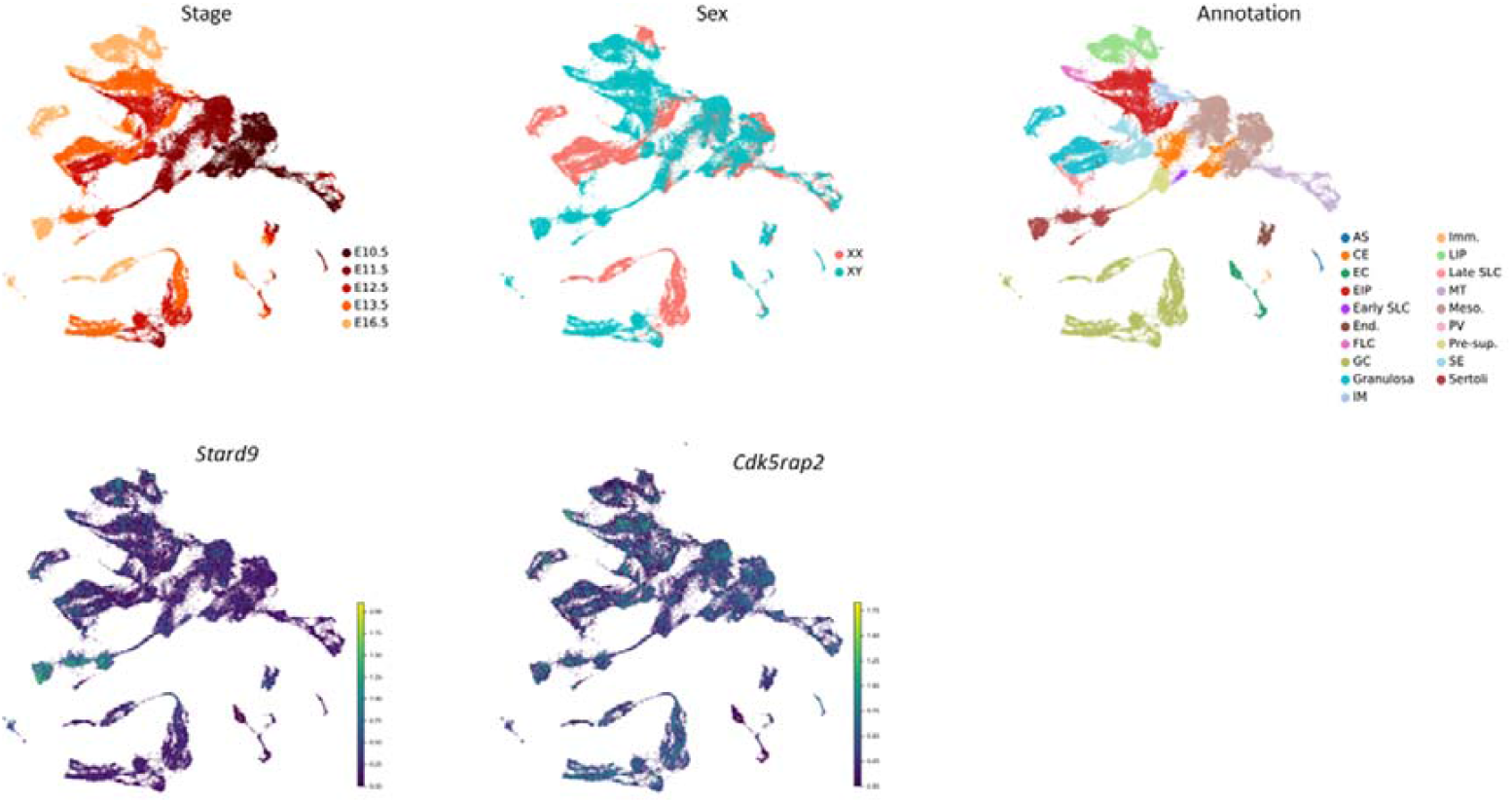
Expression of Stard9 and Cdk5rap2 in developing mouse gonads based on single-cell transcriptomics. UMAP visualization of 94,705 cells, coloured by developmental stage, sex, annotation of different cell clusters, and expression levels of *Stard9* and *Cdk5rap2* genes. Cell cluster annotations include: AS, adrenosympathic cells; CE, coelomic epithelial cells; EC, erythrocytes; EIP, early interstitial progenitors; End., endothelial cells; FLC, foetal Leydig cells; GC, germ cells; Granulosa, pregranulosa cells; IM, invading mesonephric cells; Imm., immune cells; LIP, late interstitial progenitors; MT, mesonephric tubules; Meso, mesonephric mesenchymal cells; PV, perivascular cells; Pre-sup., presupporting cells; SE, surface epithelial cells; Sertoli, Sertoli cells; SLC, supporting-like cells.

WES identified two *STARD9* variants in a compound heterozygous state in our patient. The first variant was an in-frame deletion, NM_020759.3:c.5585_5590del (p.Ser1862_Thr1863del) (rs528276071), inherited from the heterozygous father. The second variant was a missense mutation, NM_020759.3:c.3514C>T (p.Arg1172Cys) (rs12594837), inherited from the heterozygous mother. Both variants were confirmed by Sanger sequencing in the patient and her parents (**Fig. 3**).

**Fig. 3.** STARD9 and CDK5RAP2 variants identified in a patient with 46,XY complete gonadal dysgenesis (46,XY CGD). The inheritance pattern of identified variants is shown on the left side of each panel. A schematic representation of STARD9 and CDK5RAP2 proteins, highlighting functional domains and regions critical for protein-protein interactions, as well as the localization of variants identified in the patient (top-right section of each panel). Alignments of mammalian STARD9 and CDK5RAP2 fragments with arrows indicating mutation sites at the protein level (bottom-right section of each panel).

The minor allele frequency (MAF) of the p.Ser1862_Thr1863del variant is 0.001325, while the MAF of p.Arg1172Cys variant is 0.009863. Neither variant has been previously described as linked to any disease. Both lie outside functional domains of STARD9. While the Ser1862 and Thr1863 residues are conserved among mammals, the Arg1172 residue is not (**Fig. 4**).

**Fig. 4.**
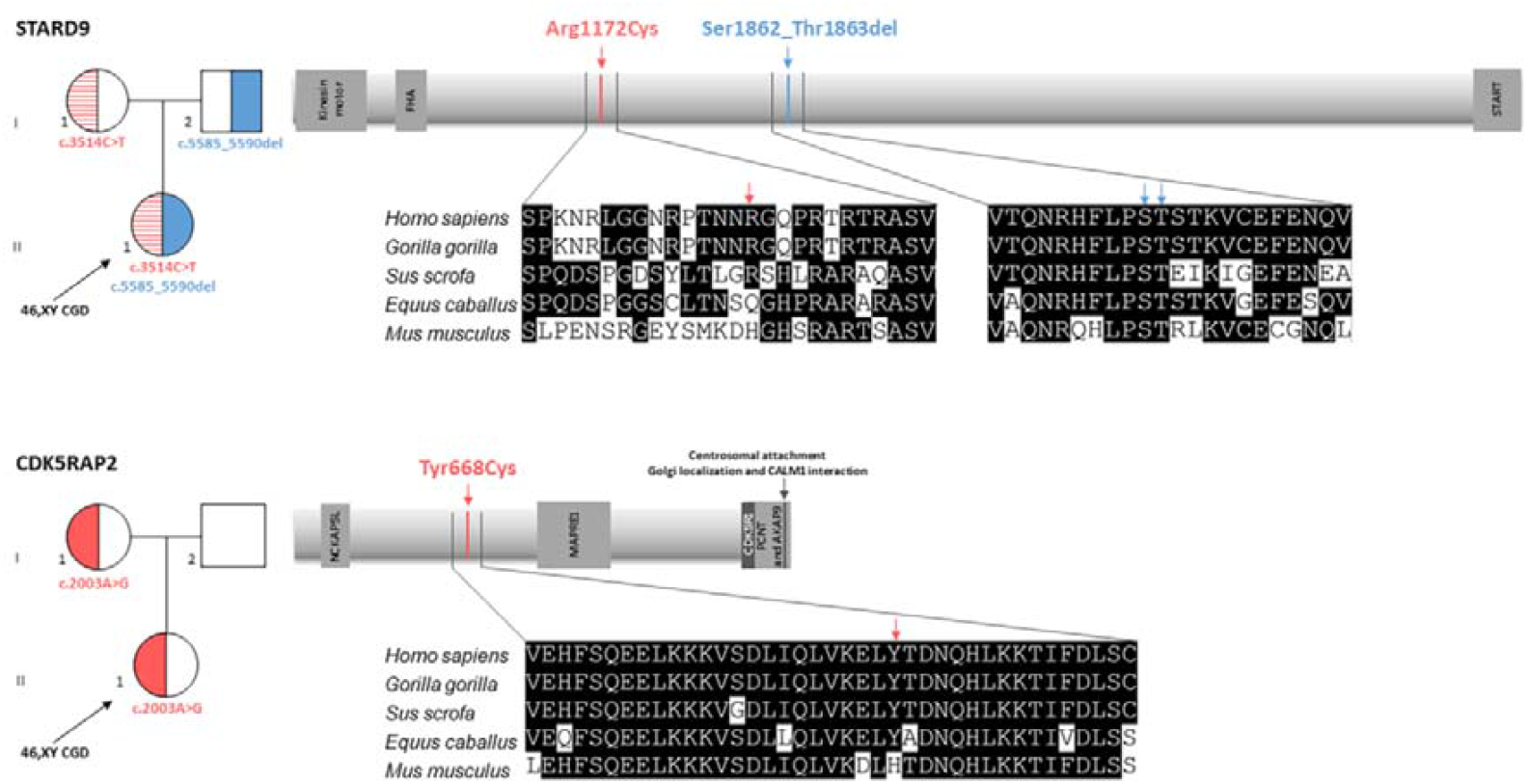
STARD9 and CDK5RAP2 variants identified in a patient with 46,XY complete gonadal dysgenesis (46,XY CGD). The inheritance pattern of identified variants is shown on the left side of each panel. A schematic representation of STARD9 and CDK5RAP2 proteins, highlighting functional domains and regions critical for protein-protein interactions, as well as the localization of variants identified in the patient (top-right section of each panel). Alignments of mammalian STARD9 and CDK5RAP2 fragments with arrows indicating mutation sites at the protein level (bottom-right section of each panel).

### Disordered nature of variant-containing regions in the STARD9 protein

To assess the potential impact of *STARD9* variants on protein structure, we applied *in silico* approaches, as the STARD9 protein has not yet been crystallized. Both identified variants, p.Arg1172Cys and p.Ser1862_Thr1863del, are located in the central region of the protein, outside functional domains. We focused on analysing this central segment to predict how these variants might alter STARD9 structure. To this end, we first predicted the secondary structure of full-length STARD9. Most of the predictors we applied indicated that Arg1172 and Ser1862_Thr1863 are located in an unstructured region (**Fig. 5a**), whereas some analyses suggested that Ser1862_Thr1863 residues could be involved in the formation of an ordered region (**Fig. 5b**). AlphaFold 3D structure predictions, further supported that these sequences are part of an intrinsically disordered region (IDR), consistent with their amino acid composition (data not shown).

**Fig. 5.**
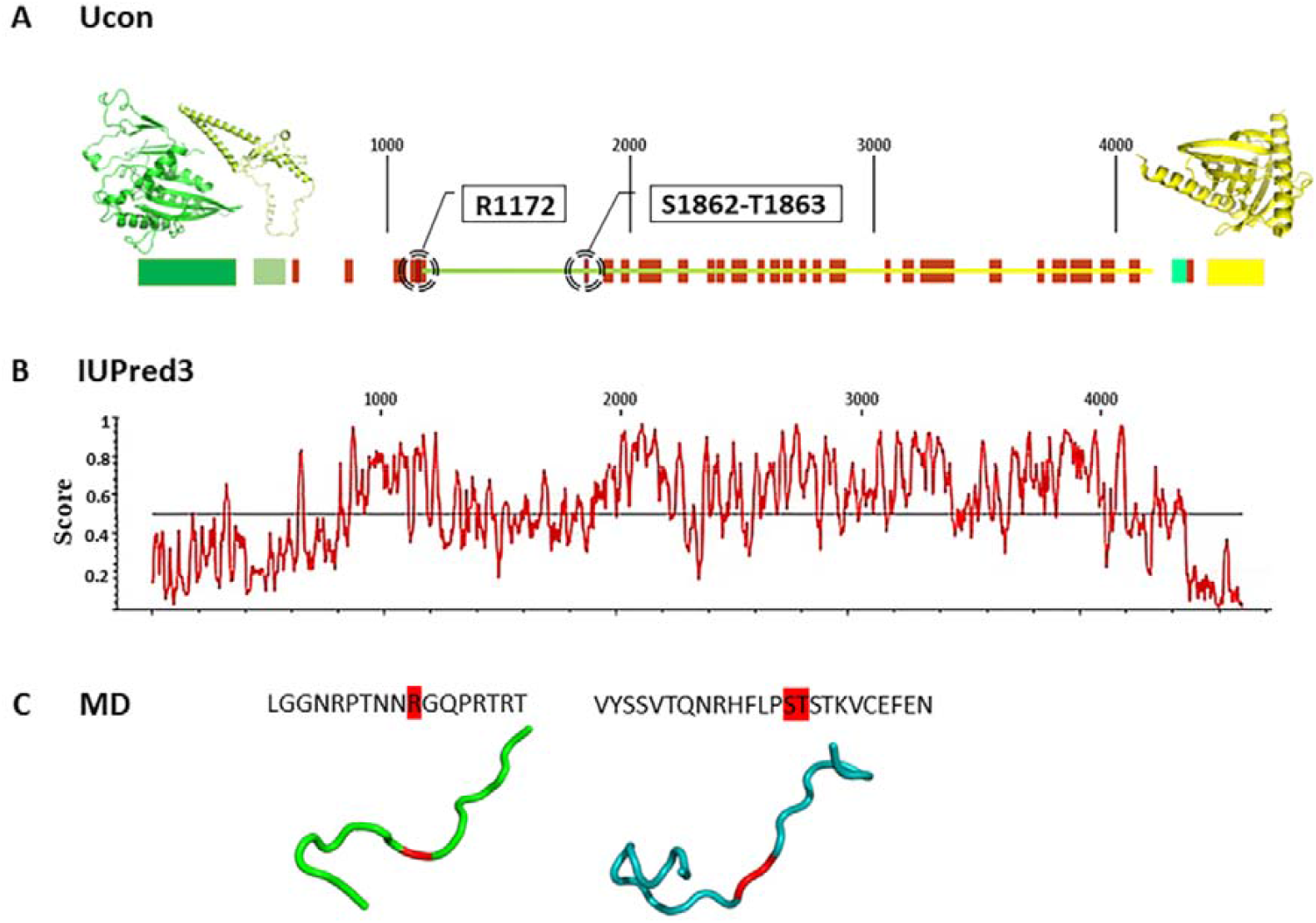
Workflow of STARD9 protein structure prediction. A. Secondary structure prediction of the full-length STARD9 protein obtained using the Ucon predictor and homology modelling to kinesin-3 KLP-6. The prediction confirmed the presence of structured domains: kinesin motor (dark green), FHA (light green), and START (yellow), and revealed many disordered regions (red). Both studied STARD9 variant sites are located within predicted disordered regions. B. Secondary structure prediction of the full-length STARD9 protein, obtained using IUPred3 predictor based on the amino acid sequence and energy estimation approach reflecting the probability of the ordered regions located upper the median and disordered regions, located lower. It predicts the region containing Arg1172 to be ordered, whereas Ser1862_Thr1863 to be disordered. C. The most frequent conformations of the short STARD9 fragments containing either the p.Arg1172Cys or p.Ser1862_Thr1863del mutation, obtained using molecular dynamics, showing the disordered structure of the studied fragments.

To validate the secondary structure predictions, we performed 3D reconstruction and MD simulations. Coarse-grain MD simulations were conducted on the wild-type STARD9 core fragment containing Arg1172 and Ser1862_Thr1863, to evaluate the behaviour in solution and assess the probable effect of these mutations. However, due to the mosaic-like structure of this protein fragment, which contained a mix of unstructured regions and secondary structure elements, we did not obtain significant results that could provide an exhaustive description of its behaviour. The fragment was highly unstable, preventing its use as a template structure for mutation analysis.

To address the limitations of coarse-grain MD simulations, we performed full-atom MD simulations on two short STARD9 mutated fragments, each containing one of the identified mutation (p.Arg1172Cys or p.Ser1862_Thr1863del). These simulations aimed to model the secondary structure folding process directly on the mutated sequences. Results revealed that both fragments retained their disordered nature, showing no tendency to form secondary structures (**Fig. 5c**).

Thus, it can be assumed that the studied mutations are most likely located in an IDR. Importantly IDRs often serve as recognition surfaces for protein-protein interactions (33, 34), suggesting that the STARD9 variants could alter interactions of that protein with its binding partners.

### A *CDK5RAP2* heterozygous variant inherited from the mother

Given that the STARD9 variants were located within a potential recognition region of the protein, and considering the oligogenic inheritance of the 46,XY CGD in our patient, we explored whether STARD9 protein interactors (**Fig. 6**) contained additional variants.

**Fig. 6.**
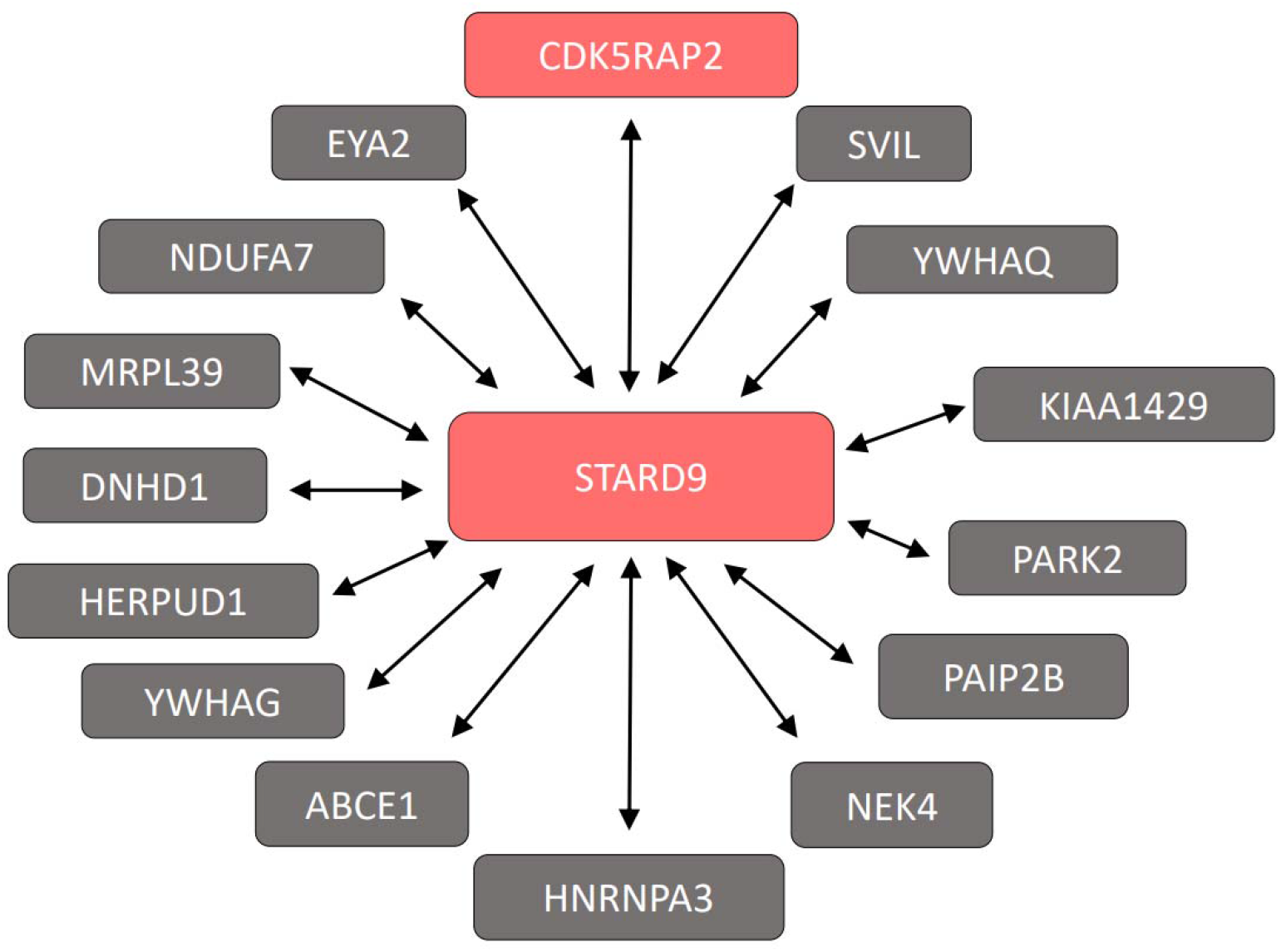
STARD9-interacting partners (based on the BioGRID and IntAct protein-protein interaction databases), expressed in the urogenital system of a fetus or adult human (according to the NCBI Gene database). Proteins highlighted in red contain rare variants detected by WES in a patient with 46,XY complete gonadal dysgenesis(46,XY CGD).

In doing so, we identified missense variant in the *CDK5RAP2* gene, which encodes a known STARD9-interacting protein (6). The identified variant, NM_018249.5:c.2003A>G (p.Tyr668Cys) (rs137966123), was inherited from the heterozygous mother (**Fig. 4**). Sanger sequencing validated the variant in both the patient and her mother (**Fig. 3**). The MAF of the *CDK5RAP2* variant is 0.0003284. The variant has not been previously described as implicated in the pathogenesis of any disease. The Tyr668 residue is not conserved and is located outside regions known to be critical for protein localization or interactions (**Fig. 6**). However, the region required for interaction with STARD9 has not been characterized to date.

### Structural analysis of wild-type CDK5RAP2 and the Tyr668Cys variant

To assess the potential structural impact of the p.Tyr668Cys variant in CDK5RAP2, we conducted *in silico* modelling using AlphaFold 2, which generated multiple structural models characterized by 9 to 14 α-helices. A central α-helix-based scaffold was a common feature across all models, forming a structural backbone of the protein. However, the smaller helices, connected by flexible linkers, exhibited variability in their spatial arrangement (data not shown).

Importantly, in all models, the p.Tyr668Cys CDK5RAP2 variant affected a central region of the α-helix (residues ∼650–684). Our 200-ns MD simulation of both the intact and mutant helices indicated a stable secondary structure in both cases. However, based on structures with similar substitutions (35, 36), this mutation may play a critical role in inter-domain interactions.

## DISCUSSION

In this study, we investigated a case of 46,XY CGD and performed WES to explore the genetic basis of the patient’s condition. No pathogenic variants were identified in genes previously associated with 46,XY GD. Instead, we discovered variants in *STARD9* and *CDK5RAP2*, two genes encoding interacting proteins. Given that 1) both genes exhibit evidence of playing a crucial role in testicular development (7, 8), and 2) recent studies indicate that some cases of 46,XY GD may follow an oligogenic inheritance pattern (3, 5), we propose that *STARD9* and *CDK5RAP2* variants together contribute to 46,XY CGD in our patient.

We propose that *STARD9* is an essential factor in testicular development for several reasons. First, *Stard9* is expressed in a sex-specific manner with the highest expression in Sertoli cells during gonadal development in mouse (7). Second, pathogenic variants in other members of the START family, such as *STAR* and *STARD8*, have been implicated in 46,XY DSD including 46,XY GD (26, 29, 32). Similarly, *CDK5RAP2* plays a critical role in gonadal development, as demonstrated in mouse models, where homozygous knockout results in underdeveloped testes and failed Sertoli cell polarization (8).

Both STARD9 and CDK5RAP2 are centrosome components that localize to the pericentriolar material (PCM) surrounding the centrioles (37, 38). STARD9 stabilizes the PCM, and its disruption leads to spindle fragmentation, apoptosis, lower microtubule coiling, and impaired chromosome segregation (38, 39). In turn, CDK5RAP2 plays a critical role in centriole replication, PCM cohesion, and microtubule nucleation and organization (37, 40).

Truncating mutations in genes encoding centrosome components, including *STARD9* and *CDK5RAP2*, have been associated with recessive microcephaly. A homozygous *STARD9* mutation resulting in a nonsense mutation in the START domain has been reported in a patient with microcephaly and dwarfism (41). Similarly, mutations in *CDK5RAP2* disrupting its centrosome localization domain lead to primary microcephaly with or without dwarfism (42). While these genes have primarily been studied in the context of brain development, their role in other organs, including the testes, has largely been neglected. Recently, a missense variant in *CDK5RAP2* was associated with non-obstructive azoospermia (43), while *STARD9* was shown to be downregulated in sperm cells of patients with asthenozoospermia (44), supporting their role in testicular function.

In our 46,XY CGD patient we identified two compound heterozygous variants in *STARD9* (p.Arg1172Cys and p.Ser1862_Thr1863del). Structural analysis revealed that these regions correspond IDRs, which are known to mediate protein post-translational modifications (PTMs) (45, 46) and protein-protein interactions (47). While little is known about the PTMs of STARD9, the residues Ser1862 and Thr1863 could potentially be phosphorylated in the wild-type, and the 1172 residue could be potentially methylated in the wild-type and/or in the p.Arg1172Cys variant. Such PTM alterations could affect STARD9’s interactions and function. Additionally, our patient carries a *CDK5RAP2* p.Tyr668Cys variant, located within an α-helix region. This variant could disrupt secondary structure contacts, alter the protein’s functional state and its interactions (36, 48, 49). The amino acid positions Arg1172 of STARD9 and Tyr668 of CDK5RAP2, exhibit a low level of conservation, which may be explained by their localization in regions of the proteins with lower packing density, as previous studies suggest such regions evolve more rapidly (46, 50).

Based on 1) the 46,XY CGD phenotype of our patient, 2) the high expression of *STARD9* and *CDK5RAP2* in Sertoli cells during gonadal differentiation (7), and 3) the role of *CDK5RAP2* in Sertoli cell polarisation (8), we propose that *STARD9* and *CDK5RAP2* variants act in concert to disrupt Sertoli cell development, leading to 46,XY CGD. To our knowledge, this is the first study associating *STARD9* and *CDK5RAP2* variants with 46,XY CGD, highlighting their potential role in early human testicular development. Based on our findings and existing evidence, we suggest that *STARD9* and *CDK5RAP2* should be included as candidate genes in genetic panels for 46,XY GD.

## DATA AVAILABILITY STATEMENT

Due to ethical restrictions, additional data are available from the corresponding author, L.L., upon reasonable request.

## ACKNOWLEDGEMENTS

We thank the family for their cooperation. We also thank our colleague, Prof. Jadwiga Jaruzelska, for reading the manuscript and providing critical comments.

## AUTHOR CONTRIBUTION STATEMENT

Conceptualization L.L., K.K.-Z.; Data curation D.S.; Formal analysis D.S., M.K., O.G.; Funding acquisition L.L., S.N.; Investigation D.S., A.R., V.K., M.K., O.S., O.G., K.K., A.K., Z.L., C.M., S.N., K.K.-Z., L.L.; Methodology D.S., K.K.-Z., L.L.; Project administration S.N., L.L.; Resources L.L.; Software D.S., A.R. ; Supervision L.L., Validation D.S., O.G., K.K., A.K.; Z.L.; Visualization D.S., V.K., C.M., K.K.-Z.; Writing - original draft D.S., A.R., K.K.-Z., L.L.; Writing - review & editing, D.S., A.R., V.K., M.K., O.S., O.G., K.K., A.K., Z.L., C.M., S.N., K.K.-Z., L.L. All authors have read and agreed to the published version of the manuscript.

## ETHICAL APPROVAL

## Study approval statement

The study was conducted according to the guidelines of the Declaration of Helsinki, and approved by the Committee on Bioethics of the Institute of Molecular Biology and Genetics of National Academy of Sciences of Ukraine (protocol code No.2, date of approval 30 April 2013).

## Consent to participate statement

Informed consent was obtained from the parents of the patient involved in the study.

## REFERENCES

1. Cools M, Nordenstrom A, Robeva R, Hall J, Westerveld P, Fluck C, et al. Caring for individuals with a difference of sex development (DSD): a Consensus Statement. Nat Rev Endocrinol. 2018;14(7):415–29.

2. Elzaiat M, McElreavey K, Bashamboo A. Genetics of 46,XY gonadal dysgenesis. Best Pract Res Clin Endocrinol Metab. 2022;36(1):101633.

3. Camats N, Fluck CE, Audi L. Oligogenic Origin of Differences of Sex Development in Humans. Int J Mol Sci. 2020;21(5).

4. Kouri C, Sommer G, Fluck CE. Oligogenic Causes of Human Differences of Sex Development: Facing the Challenge of Genetic Complexity. Horm Res Paediatr. 2023;96(2):169–79.

5. Mazen I, Abdel-Hamid M, Mekkawy M, Bignon-Topalovic J, Boudjenah R, El Gammal M, et al. Identification of NR5A1 Mutations and Possible Digenic Inheritance in 46,XY Gonadal Dysgenesis. Sex Dev. 2016;10(3):147–51.

6. Stelzl U, Worm U, Lalowski M, Haenig C, Brembeck FH, Goehler H, et al. A human protein-protein interaction network: a resource for annotating the proteome. Cell. 2005;122(6):957–68.

7. Mayere C, Regard V, Perea-Gomez A, Bunce C, Neirijnck Y, Djari C, et al. Origin, specification and differentiation of a rare supporting-like lineage in the developing mouse gonad. Sci Adv. 2022;8(21):eabm0972.

8. Kang D, Shin B, Kim GN, Hea JH, Sung YH, Rhee K. Roles of Cep215/Cdk5rap2 in establishing testicular architecture for mouse male germ cell development. FASEB J. 2024;38(22):e70188.

9. Prilusky J, Felder CE, Zeev-Ben-Mordehai T, Rydberg EH, Man O, Beckmann JS, et al. FoldIndex: a simple tool to predict whether a given protein sequence is intrinsically unfolded. Bioinformatics. 2005;21(16):3435–8.

10. Liu J, Rost B. NORSp: Predictions of long regions without regular secondary structure. Nucleic Acids Res. 2003;31(13):3833–5.

11. Linding R, Russell RB, Neduva V, Gibson TJ. GlobPlot: Exploring protein sequences for globularity and disorder. Nucleic Acids Res. 2003;31(13):3701–8.

12. Huang F, Oldfield CJ, Xue B, Hsu WL, Meng J, Liu X, et al. Improving protein order-disorder classification using charge-hydropathy plots. BMC Bioinformatics. 2014;15 Suppl 17(Suppl 17):S4.

13. Erdos G, Pajkos M, Dosztanyi Z. IUPred3: prediction of protein disorder enhanced with unambiguous experimental annotation and visualization of evolutionary conservation. Nucleic Acids Res. 2021;49(W1):W297–W303.

14. Schlessinger A, Punta M, Rost B. Natively unstructured regions in proteins identified from contact predictions. Bioinformatics. 2007;23(18):2376–84.

15. Galzitskaya OV, Garbuzynskiy SO, Lobanov MY. FoldUnfold: web server for the prediction of disordered regions in protein chain. Bioinformatics. 2006;22(23):2948–9.

16. Jumper J, Evans R, Pritzel A, Green T, Figurnov M, Ronneberger O, et al. Highly accurate protein structure prediction with AlphaFold. Nature. 2021;596(7873):583–9.

17. Abraham MJ, Murtola T, Schulz R, Páll S, Smith JC, Hess B, et al. GROMACS: High performance molecular simulations through multi-level parallelism from laptops to supercomputers. SoftwareX. 2015;1–2:19–25.

18. Huang J, MacKerell AD, Jr. CHARMM36 all-atom additive protein force field: validation based on comparison to NMR data. J Comput Chem. 2013;34(25):2135–45.

19. Machado MR, Barrera EE, Klein F, Sonora M, Silva S, Pantano S. The SIRAH 2.0 Force Field: Altius, Fortius, Citius. J Chem Theory Comput. 2019;15(4):2719–33.

20. Gorelov S, Titov A, Tolicheva O, Konevega A, Shvetsov A. DSSP in GROMACS: Tool for Defining Secondary Structures of Proteins in Trajectories. J Chem Inf Model. 2024;64(9):3593–8.

21. Arboleda VA, Lee H, Sanchez FJ, Delot EC, Sandberg DE, Grody WW, et al. Targeted massively parallel sequencing provides comprehensive genetic diagnosis for patients with disorders of sex development. Clin Genet. 2013;83(1):35–43.

22. Barseghyan H, Symon A, Zadikyan M, Almalvez M, Segura EE, Eskin A, et al. Identification of novel candidate genes for 46,XY disorders of sex development (DSD) using a C57BL/6J-Y (POS) mouse model. Biol Sex Differ. 2018;9(1):8.

23. Baxter RM, Arboleda VA, Lee H, Barseghyan H, Adam MP, Fechner PY, et al. Exome sequencing for the diagnosis of 46,XY disorders of sex development. J Clin Endocrinol Metab. 2015;100(2):E333–44.

24. Eggers S, Sadedin S, van den Bergen JA, Robevska G, Ohnesorg T, Hewitt J, et al. Disorders of sex development: insights from targeted gene sequencing of a large international patient cohort. Genome Biol. 2016;17(1):243.

25. Hughes IA, Houk C, Ahmed SF, Lee PA, Group LC, Group EC. Consensus statement on management of intersex disorders. Arch Dis Child. 2006;91(7):554–63.

26. Ilaslan E, Calvel P, Nowak D, Szarras-Czapnik M, Slowikowska-Hilczer J, Spik A, et al. A Case of Two Sisters Suffering from 46,XY Gonadal Dysgenesis and Carrying a Mutation of a Novel Candidate Sex-Determining Gene STARD8 on the X Chromosome. Sex Dev. 2018;12(4):191–5.

27. Ilaslan E, Markosyan R, Sproll P, Stevenson BJ, Sajek M, Sajek MP, et al. The FKBP4 Gene, Encoding a Regulator of the Androgen Receptor Signaling Pathway, Is a Novel Candidate Gene for Androgen Insensitivity Syndrome. Int J Mol Sci. 2020;21(21).

28. Knarston I, Ayers K, Sinclair A. Molecular mechanisms associated with 46,XX disorders of sex development. Clin Sci (Lond). 2016;130(6):421–32.

29. Sirokha D, Rayevsky A, Gorodna O, Kalynovskyi V, Zelinska N, Samson O, et al. Mutations in STARD8 (DLC3) May Cause 46,XY Gonadal Dysgenesis. Sex Dev. 2023;17(4-6):181–9.

30. Witchel SF. Disorders of sex development. Best Pract Res Clin Obstet Gynaecol. 2018;48:90–102.

31. Belanger C, Cardinal T, Leduc E, Viger RS, Pilon N. CHARGE syndrome-associated proteins FAM172A and CHD7 influence male sex determination and differentiation through transcriptional and alternative splicing mechanisms. FASEB J. 2022;36(3):e22176.

32. Finkielstain GP, Vieites A, Bergada I, Rey RA. Disorders of Sex Development of Adrenal Origin. Front Endocrinol (Lausanne). 2021;12:770782.

33. Arai M, Suetaka S, Ooka K. Dynamics and interactions of intrinsically disordered proteins. Curr Opin Struct Biol. 2024;84:102734.

34. Chakrabarti P, Chakravarty D. Intrinsically disordered proteins/regions and insight into their biomolecular interactions. Biophys Chem. 2022;283:106769.

35. Muronetz VI, Pozdyshev DV, Medvedeva MV, Sevostyanova IA. Potential Effect of Post-Transcriptional Substitutions of Tyrosine for Cysteine Residues on Transformation of Amyloidogenic Proteins. Biochemistry (Mosc). 2022;87(2):170–8.

36. Zhou W, Freed CR. Tyrosine-to-cysteine modification of human alpha-synuclein enhances protein aggregation and cellular toxicity. J Biol Chem. 2004;279(11):10128–35.

37. Barrera JA, Kao LR, Hammer RE, Seemann J, Fuchs JL, Megraw TL. CDK5RAP2 regulates centriole engagement and cohesion in mice. Dev Cell. 2010;18(6):913–26.

38. Torres JZ, Summers MK, Peterson D, Brauer MJ, Lee J, Senese S, et al. The STARD9/Kif16a kinesin associates with mitotic microtubules and regulates spindle pole assembly. Cell. 2011;147(6):1309–23.

39. Srivastava S, Panda D. A centrosomal protein STARD9 promotes microtubule stability and regulates spindle microtubule dynamics. Cell Cycle. 2018;17(16):2052–68.

40. Serna M, Zimmermann F, Vineethakumari C, Gonzalez-Rodriguez N, Llorca O, Luders J. CDK5RAP2 activates microtubule nucleator gammaTuRC by facilitating template formation and actin release. Dev Cell. 2024;59(23):3175–88 e8.

41. Okamoto N, Tsuchiya Y, Miya F, Tsunoda T, Yamashita K, Boroevich KA, et al. A novel genetic syndrome with STARD9 mutation and abnormal spindle morphology. Am J Med Genet A. 2017;173(10):2690–6.

42. Naveed M, Kazmi SK, Amin M, Asif Z, Islam U, Shahid K, et al. Comprehensive review on the molecular genetics of autosomal recessive primary microcephaly (MCPH). Genet Res (Camb). 2018;100:e7.

43. Rahimian M, Askari M, Salehi N, Riccio A, Jaafarinia M, Almadani N, et al. A novel missense variant in CDK5RAP2 associated with non-obstructive azoospermia. Taiwan J Obstet Gynecol. 2023;62(6):830–7.

44. Mao XM, Xing RW, Jing XW, Zhou QZ, Yu QF, Guo WB, et al. [Differentially expressed genes in asthenospermia: a bioinformatics-based study]. Zhonghua Nan Ke Xue. 2011;17(8):694–8.

45. Dunker AK, Brown CJ, Lawson JD, Iakoucheva LM, Obradovic Z. Intrinsic disorder and protein function. Biochemistry. 2002;41(21):6573–82.

46. Nguyen Ba AN, Yeh BJ, van Dyk D, Davidson AR, Andrews BJ, Weiss EL, et al. Proteome-wide discovery of evolutionary conserved sequences in disordered regions. Sci Signal. 2012;5(215):rs1.

47. Dunker AK, Lawson JD, Brown CJ, Williams RM, Romero P, Oh JS, et al. Intrinsically disordered protein. J Mol Graph Model. 2001;19(1):26–59.

48. Padmanabhan S, Baldwin RL. Helix-stabilizing interaction between tyrosine and leucine or valine when the spacing is i, i + 4. J Mol Biol. 1994;241(5):706–13.

49. Zondlo NJ. Aromatic-proline interactions: electronically tunable CH/pi interactions. Acc Chem Res. 2013;46(4):1039–49.

50. Gilson AI, Marshall-Christensen A, Choi JM, Shakhnovich EI. The Role of Evolutionary Selection in the Dynamics of Protein Structure Evolution. Biophys J. 2017;112(7):1350–65.

