## Supplementary information for "*STARD9* and *CDK5RAP2* – novel candidate genes for oligogenic 46,XY complete gonadal dysgenesis"

**Supplementary materials**

**Supp. List.** List of 194 genes associated with Disorders of Sexual Development (DSD).

*ADAMTS16, AKAP2, AKR1C1, AKR1C2, AKR1C4, AMH, AMHR2, ANOS1, AR, ARL6, ARX, ATF3, ATRX, B3GLCT, BBS1, BBS10, BBS12, BBS2, BBS4, BBS5, BBS7, BBS9, BCOR, BMP15, BMP4, BMP7, BNC2, CBX2, CCNQ, CDKN1C, CEP41, CHD4, CHD7, CNGA1, CREBBP, CUL4B, CUL7, CYB5A, CYP11A1, CYP11B1, CYP17A1, CYP19A1, CYP26B1, DHCR24, DHCR7, DHH, DHX37, DMRT1, DMRT2, DNMT3B, DUSP6, DUSP15, DYNC2H1, EFNB1, EPG5, ESCO2, ESPN, ESR1, ESR2, ETV4, EVC, EVC2, FAT4, FBLN2, FBXL4, FEZF1, FGF10, FGF17, FGF8, FGFR1, FGFR2, FGFR3, FIG4, FKBP4, FLNA, FLRT3, FOXL2, FRAS1, FREM2, FSHB, FSHR, GATA4, GLI3, GNRH1, GNRHR, GPC3, GRIP1, HBA1, HCCS, HDAC8, HESX1, HFE, HHAT, HNF1B, HOXA13, HOXA4, HOXB6, HS6ST1, HSD17B3, HSD17B4, HSD3B2, ICK, IL17RD, INSL3, IRF6, KISS1, KISS1R, LGR5, LEP, LEPR, LHB, LHCGR, LHFPL5, LHX3, LHX4, LHX9, LMNA, MAMLD1, MAP3K1, MCM9, MED12, MID1, MKKS, MKS1, MYBL1, NEK1, NIPAL1, NKD2, NLGN4X, NMT2, NR0B1, NR3C1, NR5A1, NSMF, OPHN1, PAX2, PCNT, PCSK1, PDE4D, PEX1, PITX2, POLR3A, POR, PROK2, PROKR2, PROP1, PSMC3IP, PTDSS1, PTK2B, PTPN11, RBBP8, RIPK4, ROR2, RSPO1, SALL1, SEMA3A, SETBP1, SMOC2, SOS1, SOX10, SOX2, SOX3, SOX9, SPECC1L, SPRY4, SRD5A2, SRY, STAR, STARD8, TAC3, TACR3, TBX15, TDRD7, TMEM70, TOE1, TOX2, TP63, TRAIP, TRIM32, TSPYL1, TTC8, TUBB3, TWIST2, UBR1, WDR11, WDR35, WDR60, WNT4, WNT5A, WNT7A, WT1, WWOX, ZEB2, ZFPM2.*

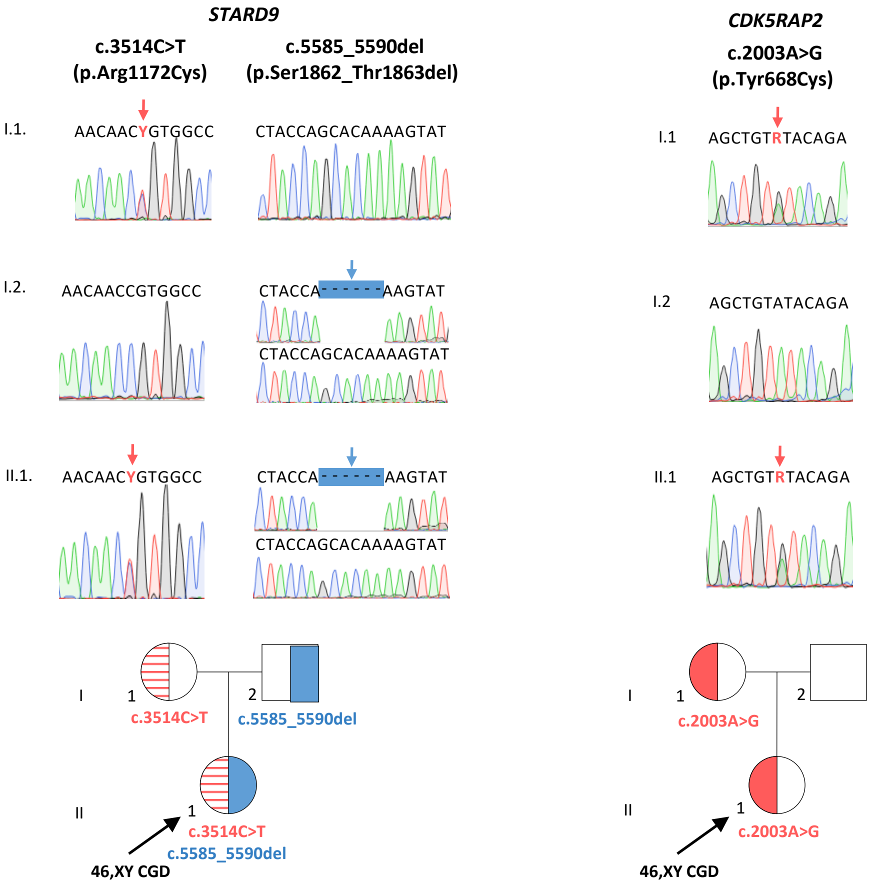


**Supp. Fig. 1.** Validation of *STARD9* and *CDK5RAP2* variants in a patient with 46,XY complete gonadal dysgenesis (46,XY CGD) using Sanger sequencing (top panel) and inheritance analysis of those variants (bottom panel).

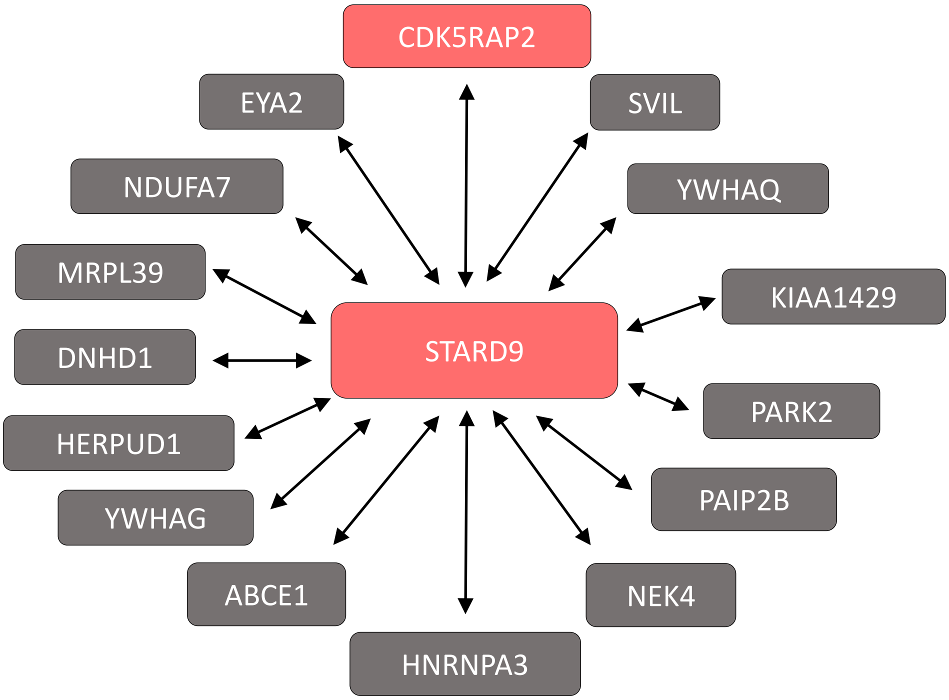


**Supp. Fig. 2.** STARD9-interacting partners (based on the BioGRID and IntAct protein-protein interaction databases), expressed in the urogenital system of a fetus or adult human (according to the NCBI Gene database). Proteins highlighted in red contain rare variants detected by WES in a patient with 46,XY complete gonadal dysgenesis (46,XY CGD).
